# Short-term associations of diarrhoeal diseases in children with temperature and precipitation in seven low- and middle-income countries from Sub-Saharan Africa and South Asia in the Global Enteric Multicenter Study

**DOI:** 10.1101/2023.12.05.23299474

**Authors:** Nasif Hossain, Lina Madaniyazi, Chris Fook Sheng Ng, Dilruba Nasrin, Xerxes Tesoro Seposo, Paul L.C. Chua, Rui Pan, Abu Syed Golam Faruque, Masahiro Hashizume

**Author notes:** **Corresponding author** Nasif Hossain, Postal address: 7-3-1 Hongo, Bunkyo-ku, Tokyo 113-0033, Japan.

## Abstract

**Background:** Diarrhoeal diseases cause a heavy burden in developing countries. Although studies have described the seasonality of diarrhoeal diseases, the association of weather variables with diarrhoeal diseases has not been well characterized in resource-limited settings where the burden remains high. We examined short-term associations between weather and hospital visits due to diarrhoea among children in seven low- and middle-income countries.

**Methodology:** Hospital visits due to diarrhoeal diseases in under 5 years old were collected from seven sites in The Gambia, Mali, Mozambique, Kenya, India, Bangladesh, and Pakistan via the Global Enteric Multicenter Study, from December 2007 to March 2011. Daily weather data during the same period were downloaded from the –ERA5-Land. We fitted time-series regression models to examine the relationships of daily diarrhoea cases with daily ambient temperature and precipitation. To account for between-site variability, we used a multivariate random-effects meta-regression model.

**Principal findings:** The relative risk (RR) of diarrhoea with temperature exposure ranged from 0.24 to 8.07, with Mozambique and Bangladesh showing positive associations, while Mali and Pakistan showing negative associations. The RR for precipitation ranged from 0.77 to 1.55, with Mali and India showing positive associations, while the only negative association was observed in Pakistan. Meta-analysis showed substantial heterogeneity in the association’s temperature–diarrhoea and precipitation– diarrhoea across sites, with *I*^*2*^ of 84.2% and 67.5%, respectively.

**Conclusions:** Child diarrhea and weather factors have diverse and complex associations across South Asia and Sub-Saharan Africa, influenced by pathogens, treatment, and nutrition. Diarrhoeal surveillance system settings should be conceptualized based on the observed pattern of climate change in these locations.

**Author Summary:** Diarrhoeal diseases remain a significant public health concern, particularly in low- and middle-income nations, and understanding the environmental factors that influence their occurrence is crucial for effective prevention and management strategies. Here, we study the weather factors, such as temperature and precipitation, that could influence the prevalence of diarrhoeal diseases in children. We found that higher temperatures were associated with an increased risk of diarrhoea in some regions. Increased precipitation was associated with a higher incidence of diarrhoea in some sites. In certain sites, diarrhoeal cases decreased with rising temperatures and precipitation. We believe that these findings may offer insights into the climate and geographic patterns of childhood diarrhoea. Such insights are crucial for developing targeted and efficient public health interventions to reduce the burden of diarrhoeal diseases. Furthermore, weather-related variables can play a significant role in driving child diarrhoea, making it imperative to identify the conditions associated with these patterns to enhance our understanding of the complex interplay between environmental factors and the prevalence of this disease.

## Introduction

Diarrhoeal disease is a major cause of death in children under 5 years old, claiming the lives of approximately 525,000 children every year [1]. Although the number of episodes of childhood diarrhoea is declining globally, the prevalence of diarrhoeal disease remains high in many resource-poor settings where infants and children are at risk of death and other infectious diseases such as malaria and pneumonia [1,2]. In South Asian regions, the diarrhoea-related morbidity remains alarmingly high, with more than 2000 children dying daily from diarrhoeal diseases, far more than the number dying from HIV/AIDS, malaria, and measles combined [3]. In Sub-Saharan Africa, diarrhoea remains a leading cause of death in children under age 5 years, accounting for approximately 750,000 of the total 4.3 million deaths among African children before their fourth birthday [4].

A complex set of weather factors is involved in driving the prevalence of diarrhoeal diseases. With climate change, there is concern that the incidence and severity of diarrhoea may increase owing to altered patterns of temperature and rainfall, as well as extreme weather events [5,6]. Temperature and rainfall variation generally impact diarrhoea prevalence through their effects on the survival and growth of various bacteria, protozoa, and viruses that cause infections of which diarrhoea is a symptom [7]. Several time-series studies have investigated the relationship between specific climatological variables and all-cause diarrhoea in particular regions, sites, and countries, including the relationship with temperature in Peru [8], Vietnam [9], China [10], Australia [11] and Fiji [12] and that with precipitation in Mozambique [13], Ecuador [14], the United States [15], and Rwanda [16]. However, most studies have focused on either temperature or rainfall, and only a few investigations have considered both the effects of temperature and precipitation on diarrhoea cases [17,18]. Additionally, these studies have been limited to analysing weekly or monthly diarrhoea counts, and none have precisely quantified the association between temperature and precipitation with daily diarrhoea counts in multiple settings and diverse contexts. The objective of this study was to investigate the short-term relationship of childhood diarrhoea with temperature and precipitation in seven low- and middle-income countries by age group. We analysed the data from the Global Enteric Multicenter Study (GEMS). The GEMS is the largest, most comprehensive study of childhood diarrhoeal diseases ever conducted in developing country settings [19].

## Methods

### Data collection

Hospital visits due to diarrhoeal diseases among children under 5 years of age were collected from the GEMS between December 1, 2007 and March 3, 2011. We included patients aged 0–59 months who met the case definition for diarrhoea (≥3 abnormally loose stools within 24 hours) and were referred to as having all-cause diarrhoea (ACD). Using dates on patients’ individual records, we generated a site-by-site time-series dataset. In the absence of an individual record on a given day, we assumed no patients on that day [20]. This study included seven study sites across seven countries: Basse (The Gambia), Bamako (Mali), Nyanza Province (Kenya), Manhiça (Mozambique), Kolkata (India), Tangail (Bangladesh), and Karachi (Pakistan). These seven sites provide good coverage for child populations in Sub-Saharan Africa and South Asia, where approximately 90% of diarrhoea mortality had been documented [21].

Hourly weather data over the study period were from the European Centre for Medium-Range Weather Forecast Reanalysis Version 5 - Land (ERA5-Land) and downloaded from the Copernicus Climate Data Store (https://cds.climate.copernicus.eu/#!/home). The dataset from ERA5-Land provides high spatial and temporal resolution, allowing for 9 km grid at hourly time steps; vertical coverage is from 2 metres above the surface level. These datasets include hourly 2-metre temperature (degree Kelvin), hourly dew point temperature (degree Kelvin), and hourly precipitation (metres). The hourly time-series variables were converted into local time zones (S1 Table) from Coordinated Universal Time (UTC)±00:00. We calculated the daily averages for temperature and dew point temperature (converted into °C) and aggregated daily total precipitation in millimetres.

The weather data were matched with the daily patient count for ACD at each site, based on location (nearest ERA5 grid square centre of each study site) and date. Our initial inspection of the data suggested highly skewed precipitation data (S2 Fig, S1 Table). Thus, we calculated the 21-day rolling sum of daily total precipitation for risk assessment [22–24]. Relative humidity was calculated as percentage using temperature and dew point temperature [20].

### Statistical analysis

To assess the associations of temperature and precipitation with daily diarrhoea visits in children, we fitted a time-series regression model with quasi-Poisson distribution and distributed lag nonlinear model for each country:

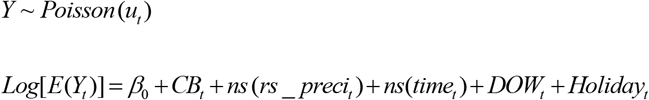

Where *Y*_*t*_ is the number of diarrhoea cases on day *t* for the diarrhoea outcome; *β*_0_ is the intercept; *CB* is the cross-basis function of temperature (smoothed using a natural cubic spline [*ns*] with 3 degrees of freedom [df]) and its lags (21-day lags with 3 equally spaced knots in the log scale). *ns(rs_preci)* is a natural cubic spline (ns) of the 21-day rolling sum of precipitation (rs_preci) with 3 df included as another main exposure; *ns(time)* is a natural cubic spline of day with 7 df per year to control for seasonality and long-term trends. *DOW*_*t*_ is an indicator variable for the day of the week on day *t*, and *Holiday*_*t*_ is an indicator variable for country-specific national holidays on day *t*. Using the same approach, we conducted additional analyses by stratifying into age groups of 0–11 months, 12–23 months, and 24–59 months. The overall cumulative relative risk (RR) for both temperature and precipitation was estimated as the ratio of the risk at the 95^th^ percentile compared to the risk at the 1^st^ percentile of each variable.

We applied a multivariate random-effects meta-analysis to assess heterogeneity and subsequently, a meta-regression to identify location-specific meta-predictors that could account for the observed heterogeneity [26]. We included site-specific average temperature, total precipitation (annual average), gross domestic product per capita, population density, poverty level, use of treated water, primary source of drinking water, handwashing practice, nutritional status (stunting), viruses, bacteria, protozoa, moderate to severe signs of dehydration, and use of oral rehydration solution (ORS) into the model as meta-predictors (S3 Table). We assessed heterogeneity using the *I*^*2*^ statistic, Cochran Q test, and likelihood ratio tests for the meta-predictors.

### Sensitivity analysis

Several sensitivity analyses were performed to test the robustness of our results. We fitted our models to control for relative humidity and dew point temperature. We changed the lag period for temperature (14 and 28 days), the duration of rolling sum for precipitation (14 and 28 days), the df for temperature (4 to 5), and the df for the 21-day rolling sum for precipitation (4 to 5).

All statistical analyses were conducted with R software version 4.2.0 (The R Foundation for Statistical Computing, Vienna, Austria). The *dlnm, mixmeta and ncdf4* packages ware used for nonlinear model with distributed lags, meta regression and ERA5 data extraction [26,27].

## Results

### Description of diarrhoea and weather factors

Table 1 shows summary statistics for the outcomes by age group and exposure variables in each country. The analysis included 66,056 ACD cases in total, with the most (n=21,751, 33%) cases in Pakistan and the fewest (n=2384, 4%) in Kenya. Diarrhoea was more common among 0–11 months (n=28,392, 43%) than in 12–23 months (n=22,487, 34%) and 24–59 months (n=15,177, 23%). Daily temperatures ranged from 15.2°C in Bangladesh to 37.7°C in Mali over the study period. The temperature variability in the locations from South Asia was comparable to that in Sub-Saharan Africa. Furthermore, the temperature distribution was notably narrow in Kenya and the widest in Pakistan (Table S1). Childhood diarrhoea showed clear seasonality in most countries (S1 Fig); however, these patterns varied across different geographic locations (S1 Fig).

**Table 1.** Summary Statistics of Daily Study Variables in Seven Sites, 2007–2011.

| Study Site | All-cause diarrhoea, n (%) |  |  |  | Average Temperature °C (range) | Total Precipitation mm (range) |
| --- | --- | --- | --- | --- | --- | --- |
|  | Age 0–11 months | Age 12–23 months | Age 24–59 months | Total |  |  |
| The Gambia | 3047 (40%) | 3415 (44%) | 1226 (16%) | 7688 | 28.8 (21.1, 37.4) | 5.48 (0.0, 305.7) |
| Mali | 4315 (47%) | 3132 (34%) | 1791 (19%) | 9238 | 27.5 (17.4, 37.7) | 6.90 (0.0, 135.7) |
| Mozambique | 5488 (42%) | 4914 (38%) | 2583 (20%) | 12985 | 23.5 (16.0, 32.5) | 4.89 (0.0, 145.7) |
| Kenya | 1148 (48%) | 706 (30%) | 530 (22%) | 2384 | 22.0 (18.8, 25.3) | 13.4 (0.0, 118.5) |
| India | 1967 (41%) | 1716 (36%) | 1085 (23%) | 4768 | 26.4 (15.9, 33.3) | 11.4 (0.0, 209.8) |
| Bangladesh | 3410<br>(47%) | 2505<br>(35%) | 1327<br>(18%) | 7242 | 25.5 (15.2, 31.9) | 15.6 (0.0, 326.2) |
| Pakistan | 9017<br>(41%) | 6099<br>(28%) | 6635<br>(31%) | 21751 | 26.2 (15.9, 34.1) | 1.5 (0.0, 132.5) |

### Association between temperature and diarrhoea

Fig 1 and Table 2 show the cumulative effect of temperature on ACD. The association between temperature and ACD tended to be linear in most countries, although the ACD risk in Mali and Bangladesh levelled off when temperatures exceeded 26°C. The RR of diarrhoea increased in Mozambique (4.96, 95% confidence interval [CI] 2.81–8.76) and Bangladesh (8.07, 95% CI 3.70– 17.60). However, the RR decreased in Mali (0.25, 95% CI 0.11–0.55) and Pakistan (0.24, 95% CI 0.12–0.51). We plotted the distributed lag for a maximum of 21 days and age-specific ACD each site (supplemental). At the 95^th^ percentile temperature, the largest effect estimates were observed on the current day (lag 0) in all seven countries, and a positive effect was observed up to 21 days in Mozambique, Kenya, and Bangladesh (S3 Fig). A significant positive association of ACD and temperature (at the 95^th^ percentile) was found at lag 0–1 in The Gambia, at lag 4–18 in Mozambique, at lag 10–16 in Kenya, and at lag 4–9 in Bangladesh. A significant negative association of ACD and temperature (at the 95^th^ percentile) was found at lag 3, lag 10–15 in Mali, and at lags 2–3 and 15–19 in Pakistan. In Bangladesh, with temperatures above the 95^th^ percentile, the cumulative RRs were higher among infants (aged 0–11 months) than in toddlers (aged 12–23 months) and children (aged 24–59 months). Conversely, in Mozambique, RRs were higher in toddlers compared with those in infants and children, although the CIs overlapped among these age groups in both countries (S4 Fig and S2 Table).

**Table 2.** Relative Risk of Temperature and Precipitation in All-Cause Diarrhoea in Seven Countries.

| Country | Temperature |  |  | Precipitation |  |  |
| --- | --- | --- | --- | --- | --- | --- |
|  | 1st pctl | 95th pctl | Relative Risk (95% CI) | 1st pctl | 95th pctl | Relative Risk (95% CI) |
| The Gambia | 23 | 34 | 1.67 (0.89, 3.14) | 0 | 535 | 1.36 (0.93, 1.99) |
| Mali | 20 | 33 | **0.25 (0.11, 0.55)** | 0 | 542 | **1.51 (1.03, 2.20)** |
| Mozambique | 17 | 29 | **4.96 (2.81, 8.76)** | 6 | 306 | 0.89 (0.73, 1.08) |
| Kenya | 20 | 24 | 2.05 (0.75, 5.58) | 57 | 550 | 1.13 (0.71, 1.81) |
| India | 17 | 32 | 0.83 (0.23, 2.94) | 0 | 769 | **1.55 (1.08, 2.22)** |
| Bangladesh | 17 | 30 | **8.07 (3.70, 17.60)** | 0 | 1011 | 1.18 (0.87, 1.61) |
| Pakistan | 17 | 31 | **0.24 (0.12, 0.51)** | 0 | 161 | **0.77 (0.67, 0.89)** |

We used a cross-basis function for temperature with a 21-day lag period both smoothed using a natural cubic spline with 3 degrees of freedom. Precipitation is based on the rolling sum of 21 days smooth using a natural cubic spline with 3 degrees of freedom. pctl, percentile; CI, confidence interval.

**Fig 1.**
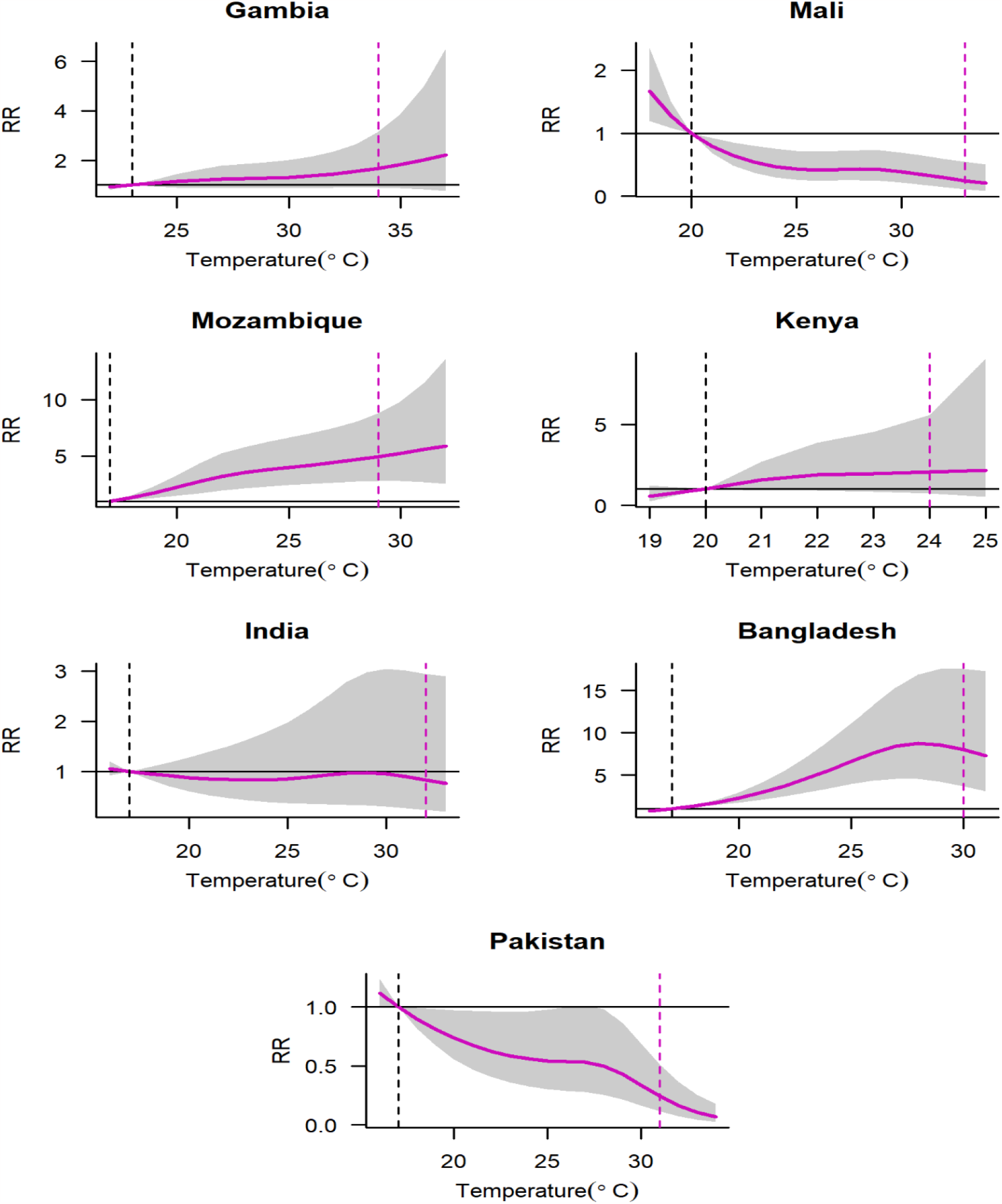
Association between daily mean temperature over 21 days and all-cause diarrhoea. Purple solid lines represent point estimates of relative risk (RR); grey shaded polygons are 95% confidence intervals. Dashed black vertical lines are reference temperature at the 1st percentile and dashed purple vertical lines are temperature at 95^th^ percentile.

**Fig 2.**
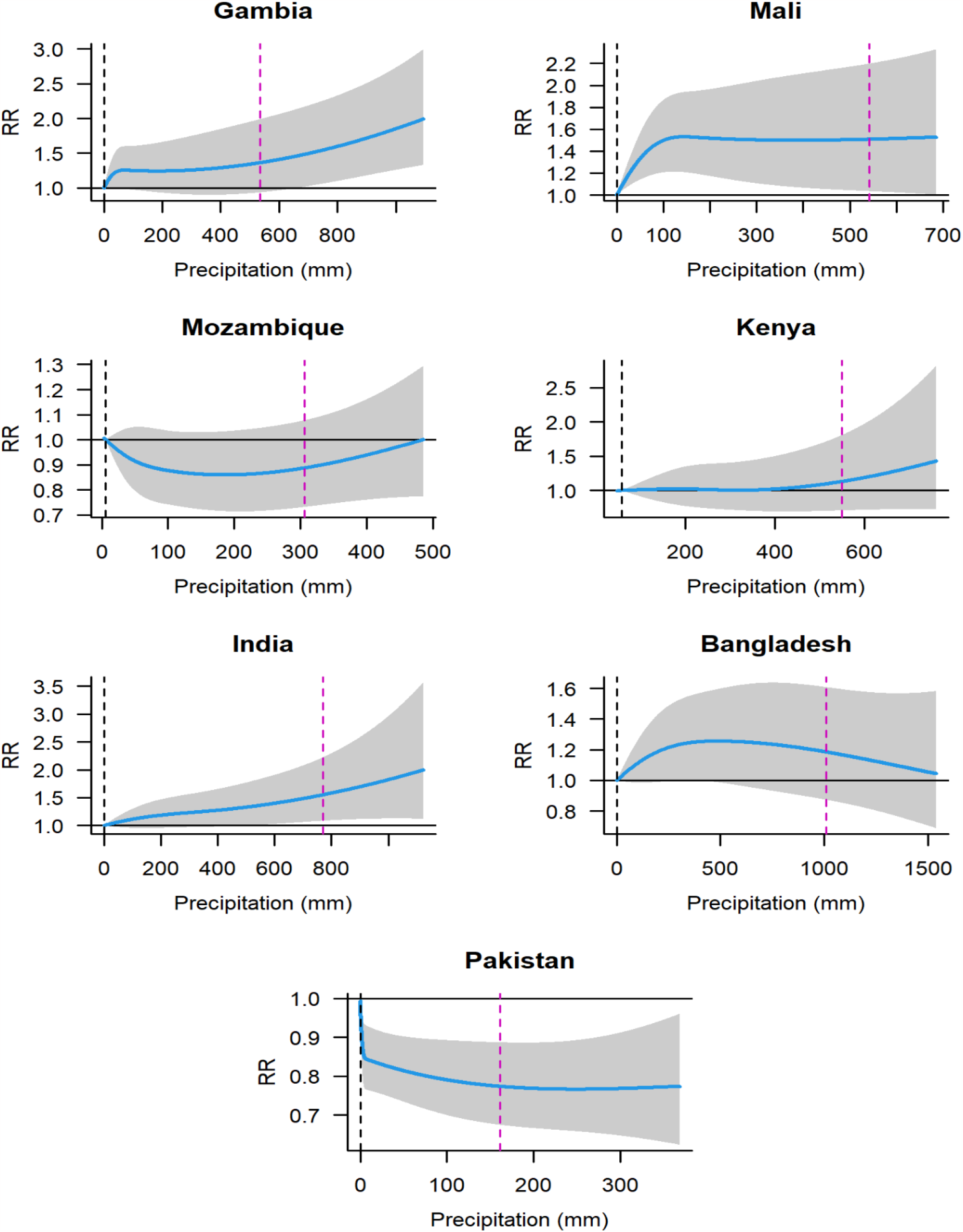
Association between precipitation over 21 days and all-cause diarrhoea. Blue solid lines represent point estimates of relative risk (RR); grey shaded polygons are 95% confidence intervals. Dashed black vertical lines are reference precipitation at the 1st percentile and dashed purple vertical lines are precipitation at the 95^th^ percentile.

### Association between precipitation and diarrhoea

The association between precipitation and ACD infection exhibited a monotonic increasing pattern in Gambia, Mali, Kenya, and India; like temperature Mali and Bangladesh also levelled off at higher precipitations. Pakistan also levelled off when rolling sum precipitation exceeded 30 mm but in the opposite direction. The RR significantly increased in Mali (1.51, 95% CI 1.03–2.20)) and India (1.55, 95% CI 1.08–2.22). The RR declined significantly (0.77, 95% CI 0.67–0.89) in Pakistan. In Mali and India, both 12-23 and 24-59 in both countries tended to exhibit higher risk than the youngest 0-11 category. However, there were overlapping CIs among these age groups in both countries (S5 Fig and S2 Table).

The simple meta-analysis showed substantial heterogeneity in the associations of temperature– diarrhoea and precipitation–diarrhoea across sites, with an *I*^*2*^ of 84.2% and 67.5%, respectively. Results of meta-regression showed that pathogens (bacteria) and dehydration treated with ORS could modify the temperature–diarrhoea association across different study sites. Only nutritional status (stunting) was found to be important predictor that modified the precipitation–diarrhoea association (Table 3).

**Table 3.**
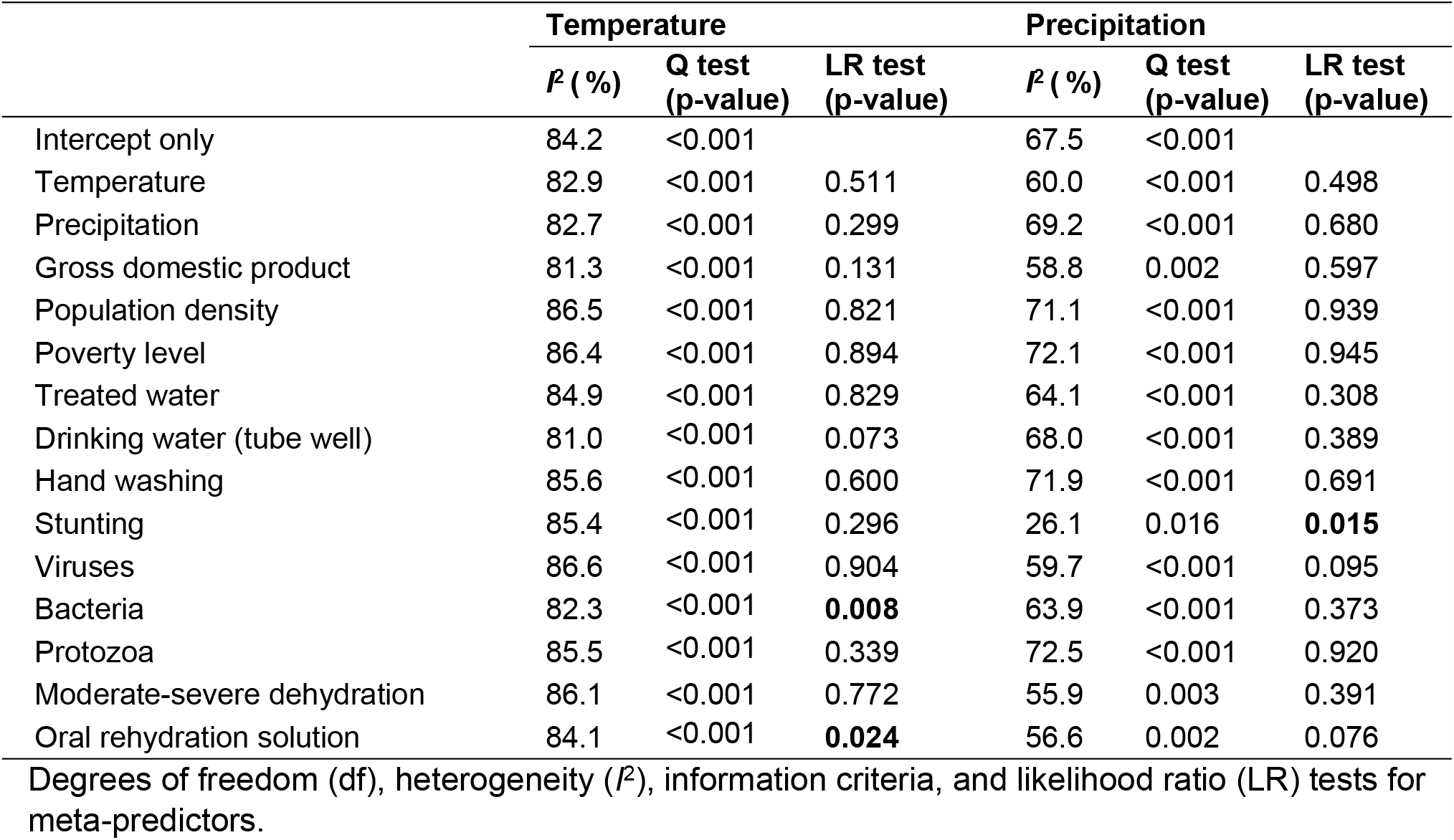
Results of meta-regression model for temperature–diarrhoea and precipitation– diarrhoea.

### Sensitivity analysis

In sensitivity analyses, the estimates for the association of temperature and precipitation with diarrhoea were generally similar when adjusting for possible time-varying covariates (relative humidity and dew point). The RRs were not substantially changed by changing the lag days, number of rolling sum days, or changing the exposure df; only some estimated CIs were reduced or increased (S6 Fig and S7 Fig).

## Discussion

We found varying patterns of diarrhoea morbidity in relation to temperature and precipitation across sites in this study. Specifically, we observed independent increase of diarrhoea morbidity associated with high temperatures in Mozambique and Bangladesh. In contrast, we found a significant inverse association between temperature and diarrhoea incidence in Mali and Pakistan. Regarding precipitation, the results demonstrated that high precipitation was independently associated with increased diarrhoea in Mali and India whereas a negative association was found in Pakistan.

The relationship between temperature and ACD varied among countries. In Mozambique and Bangladesh, the evidence indicated that high temperatures contributed to ACD morbidity. This was consistent with the findings of previous single-site studies [8,10,11,17,28–30]. A study in Mozambique found that each 1°C increase on the hottest day of the concurrent week was associated with a 3.64% increase in the incidence of diarrhoea [13]. Another study in Dhaka, Bangladesh found that an average 1°C increase in temperature over the previous 4 weeks was associated with a 6.5% increased incidence of non-cholera and non-rotavirus diarrhoea [18]. However, it is important to note that the direction of this association varies by region. In Mali and Pakistan, we found that diarrhoea morbidity decreased with increasing temperature. A few epidemiological studies have suggested that the risk of infectious diarrhoea decreases with a rise in temperature; a time-series study conducted by Thiam et al. in 2017 showed that nighttime temperatures above 26°C were negatively associated with under-five diarrhoea incidence [31], Wang et al. reported an association between meteorological factors and diarrhoea incidence in southern China for 11 years of time-series studies and found a high overall cumulative RR for low mean temperature [32]. Another study found in Botswana a negative association between temperature and diarrhoea during the wet season [33]. The results of multivariate meta-regression showed that some modifiers could explain heterogeneity among seven sites in seven countries. We found that the prevalence of bacterial pathogens modified the association between high temperature and child diarrhoea. A systematic review and meta-analysis study showed significant differences of association with temperature among bacterial pathogens [34]. The majority of studies reported positive associations between temperature and bacterial infections [35] and a study showed a negative association [36]. We also found that ORS used at home for treating diarrhoea had a significantly different effect on the association between temperature and diarrhoea. Studies have shown contrasting patterns of ORS adoption in Kenya [37] and Mexico. These modifiers reflected the differences in the local epidemiology and health behaviors of the seven sites in seven countries. These results indicate that temperature may have varying impacts on diarrhoeal diseases in different settings.

There was a monotonic increase in precipitation-related diarrhoea, but this varied in direction across locations. A steady rise in ACD cases was observed in The Gambia, Mali, Kenya, and India, with Mali and India having significant results of 542 mm and 769 mm of rainfall over a 3-week moving sum. Our findings are consistent with those of previous studies conducted in different regions [38–40]. For example, Carlton et al. performed a study in Ecuador and found that heavy rainfall events were associated with increased diarrhoea incidence following dry periods and a decrease following wet periods [41]. A study in India showed that heavy precipitation can cause water contamination and disrupt sanitation systems, with a breakdown of infrastructure [28]. Additionally, heavy precipitation is associated with an increased risk of diarrhoea through its impact on river levels [42], causing river levels to rise, which can lead to flooding. Flooding can lead to contamination of water sources with sewage and other pollutants, making it difficult access clean water and increasing the risk of waterborne illnesses such as diarrhoea. Heavy precipitation can also cause a flushing action that can send pathogens, including those that cause diarrhoea, into surface and groundwater sources. This can happen when heavy precipitation causes surface runoff, which can carry bacterial and protozoan pathogens from contaminated areas [43] such as from overflowing septic tanks, latrines, or sewage treatment plants, into nearby water sources [44]. In contrast, our analysis revealed an inverse relationship between precipitation and diarrhoea in Pakistan, which receives the lowest amount of rain among the countries in our study (see Table 1). This implies that low precipitation levels increase the risk of diarrhoea in children; previous studies support these findings [16,29]. A lack of water owing to drought has been associated with outbreaks of several waterborne diseases. For instance, in places where water is scarce, there has been an increased occurrence of infections like cholera, *Escherichia coli* infection, and leptospirosis [45], which could be attributed to poor hygiene practices and the consumption of contaminated water with higher pathogen concentrations [6]. However, the association between precipitation and diarrhoea differed across different sites in our study. We investigated this heterogeneity and found that nutritional status (stunting) could modify this association. It is less clear how to interpret these variations. A study investigated the association between rainfall variations and child diarrhoea in 15 Sub-Saharan African countries and shows that height for age z score is no longer significant by climate zones [7]. Nevertheless, a study showed watery diarrhoea was 63% less likely to cause death in children with acute malnutrition than those with better nutritional status [46].

Our age-stratified analysis revealed that children under two years of age were more susceptible to diarrheal diseases in relation to climate change. This could be explained by the fact that very young children may stay indoors with their mothers, who are often engaged in domestic activities that expose them to contaminated water and food sources [47]. The immune systems of young children may ware not fully developed and may not be able to cope with the increased pathogen load that results from climate change [48]. Moreover, when they start to walk, they may encounter more environmental exposure like temperature and precipitations that could increase their risk of infection [49].

The relationship among temperature, precipitation, and diarrhoea is complex and can vary depending on the specific pathogens involved. Different pathogens have different transmission mechanisms and survival rates under different environmental conditions. For example, some diarrhoeal pathogens may survive longer in warmer temperatures whereas others may thrive in wetter conditions. Therefore, conducting pathogen-specific analysis in future studies is crucial to better understand the relationship between environmental factors and diarrhoeal diseases.

### Strengths and limitations of this study

Several studies have investigated the association between diarrhoea and environmental factors, such as temperature and precipitation, in one country or multiple sites in one country. However, there is little research on this association across multiple countries using consistent methods. We addressed this knowledge gap by analysing data from multiple site and geographically varied countries using well-tested methods to investigate the association between diarrhoea and environmental factors, such as temperature and precipitation. The daily dataset, which enables the application of advanced DLNM to assess delayed effects, it would be beneficial to elaborate on this in the discussion section in the context of temperature. This is especially since the results section has distinctly outlined the lag patterns across various sites. The seven study sites represent a range of child health indicators in developing countries as well as urban, peri-urban, and rural settings. The study performed an analysis stratified by age, which is a rare approach taken in similar topics. To our best knowledge, we believe that this one is the largest study addressing the independent impact of temperature and precipitation on ACD, with consideration of non-linear and delayed dependencies. We examined the associations of diarrhoea with temperature and precipitation in terms of total as well as age-specific diarrhoea morbidity.

Some study limitations must be acknowledged. First, we only considered one site in each country; therefore, our findings may not be nationally representative. Second, the study period was relatively short, and CIs of the estimates were wide for some sites. Lastly, pathogens causing diarrhoea infections might behave differently under different climate conditions, and our dataset failed to build up the DLNM model due to the probability of daily sampling is not equal. Future research should focus on the mechanism of temperature and precipitation on diarrhoeal infectious diseases, such as in pathogen-specific analysis and projection of future risks of diarrhoea considering socioeconomic pathways.

## Conclusion

Our summary findings suggested an essential role of both ambient temperature and precipitation in increasing the likelihood of diarrhoeal infections in South Asian and Sub-Saharan regions. Specifically, Mozambique, and Bangladesh were found to be more susceptible to diarrhoeal diseases with high temperatures whereas Mali and Pakistan exhibited a decrease in infections. Similarly, high precipitation was linked to an increase in diarrhoeal infections in Mali and India but a decrease in infections in Pakistan. It is possible that this variation can be explained by factors such as disease-causing pathogens, the use of ORS for treating dehydration, and nutritional status.

## Data Availability

No - some restrictions will apply

https://docs.google.com/document/d/1okWceRFqS7QGZcveKWJ5NX1s1MhITkbY/edit?usp=sharing&ouid=110974106304595297238&rtpof=true&sd=true

## Funding

This work was supported by the Japan Science and Technology Agency (JST) as part of SICORP, Grant Number JPMJSC20E4, and the Japanese Government (Monbukagakusho) Scholarship from the Ministry of Education, Science, Sport, and Culture of Japan.

## Acknowledgements

We gratefully acknowledge the support of the University of Maryland, which is the PI Institute of GEMS study. We thank Toshihiko Sunahara, PhD, and Michiko Toizumi, PhD for editing a draft of this manuscript.

## Author Contributions

Conceptualization: Nasif Hossain, Masahiro Hashizume

Data curation: Nasif Hossain

Formal analysis: Nasif Hossain

Investigation: Nasif Hossain

Methodology: Nasif Hossain, Lina Madaniyazi, Chris Fook Sheng Ng, Masahiro Hashizume

Supervision: Lina Madaniyazi, Chris Fook Sheng Ng, Dilruba Nasrin, Masahiro Hashizume

Project administration: Masahiro Hashizume

Resources: Xerxes Tesoro Seposo, Paul L.C. Chua, Rui Pan, Abu Syed Golam Faruque

Software: Masahiro Hashizume

Validation: Nasif Hossain

Visualization: Nasif Hossain

Writing – original draft: Nasif Hossain

Writing – review & editing: Nasif Hossain, Lina Madaniyazi, Chris Fook Sheng Ng, Dilruba Nasrin, Xerxes Tesoro Seposo, Paul L.C. Chua, Rui Pan, Abu Syed Golam Faruque, Masahiro Hashizume

Funding acquisition: Nasif Hossain, Masahiro Hashizume

## Supporting information

**S1 Fig. Time-series plots of the daily number of all-cause diarrhoea cases, moderate to severe diarrhoea cases, daily mean temperature, and precipitation between 2008 and 2011 in the study countries**.

(DOCX)

**S2 Fig. Distribution of ERA5 daily temperature and precipitation**.

(DOCX)

**S3 Fig. Lag-response relationship between temperature and ACD morbidity at 95**^**th**^ **percentile temperature for each site**.

(DOCX)

**S4 Fig. Association between daily mean temperature over 21 days and all-cause diarrhoea stratified by age group**.

(DOCX)

**S5 Fig. Association between daily precipitation over 21 days and all-cause diarrhoea stratified by age group**.

(DOCX)

**S6 Fig. Cumulative relative risks (RRs) with 95% Cis [95**^**th**^ **vs. 1**^**st**^ **Pctl] by country in sensitivity analyses for the Temperature-Diarrhoea model**.

(DOCX)

**S7 Fig. Cumulative relative risks (RRs) with 95% Cis [95**^**th**^ **vs. 1**^**st**^**] by country in sensitivity analyses for the Precipitation-Diarrhoea model**.

(DOCX)

**S1 Table. Summary statistics information of ERA5 land**

(DOCX)

**S2 Table. Summary of age-specific relative risk of temperature (°C) and precipitation over 21 days on all-cause diarrhoea for seven countries at 95**^**th**^ **percentile temperature and precipitation**.

(DOCX)

**S3 Table S3. List of site-specific meta predictors**

(DOCX)

## Notes

### Competing Interest Statement

The authors have declared no competing interest.

### Funding Statement

Yes

### Author Declarations

We have taken ethical approval from School of Tropical Medicine and Global Health (TMGH), Nagasaki University.

